# Associations between SARS-CoV-2 infection and subsequent economic inactivity and employment status: pooled analyses of five linked longitudinal surveys

**DOI:** 10.1101/2023.07.31.23293422

**Authors:** Richard J Shaw, Rebecca Rhead, Richard J Silverwood, Jacques Wels, Jingmin Zhu, Olivia KL Hamilton, Giorgio Di Gessa, Ruth CE Bowyer, Bettina Moltrecht, Michael J Green, Evangelia Demou, Serena Pattaro, Paola Zaninotto, Andy Boyd, Felix Greaves, Nish Chaturvedi, George B. Ploubidis, Srinivasa Vittal Katikireddi

## Abstract

**Introduction:** Following the acute phase of the COVID-19 pandemic, record numbers of people became economically inactive (i.e., neither working nor looking for work), or non-employed (including unemployed job seekers and economically inactive people). A possible explanation is people leaving the workforce after contracting COVID-19. We investigated whether testing positive for SARS-CoV-2 is related to subsequent economic inactivity and non-employment, among people employed pre-pandemic.

**Methods:** The data came from five UK longitudinal population studies held by both the UK Longitudinal Linkage Collaboration (UK LLC; primary analyses) and the UK Data Service (UKDS; secondary analyses). We pooled data from five long established studies (1970 British Cohort Study, English Longitudinal Study of Ageing, 1958 National Child Development Study, Next Steps, and Understanding Society). The study population were aged 25-65 years between March 2020 to March 2021 and employed pre-pandemic. Outcomes were economic inactivity and non-employment measured at the time of the last follow-up survey (November 2020 to March 2021, depending on study). For the UK LLC sample (n=8,174), COVID-19 infection was indicated by a positive SARS-CoV-2 test in NHS England records. For the UKDS sample we used self-reported measures of COVID-19 infection (n=13,881). Logistic regression models estimated odds ratios (ORs) with 95% confidence intervals (95%CIs) adjusting for potential confounders including sociodemographic variables, pre-pandemic health and occupational class.

**Rebsults:** Testing positive for SARS-CoV-2 was very weakly associated with economic inactivity (OR 1.08 95%CI 0.68-1.73) and non-employment status (OR 1.09. 95%CI 0.77-1.55) in the primary analyses. In secondary analyses, self-reported test-confirmed COVID-19 was not associated with either economic inactivity (OR 1.01 95%CI 0.70-1.44) or non-employment status (OR 1.03 95%CI 0.79-1.35).

**Conclusions:** Among people employed pre-pandemic, testing positive for SARS-CoV-2 was either weakly or not associated with increased economic inactivity or non-employment. Research on the recent increases in economic inactivity should focus on other potential causes.

**Key messages:** *What is already known on this topic:*

- Economic inactivity has increased following the pandemic.
- Infection with SARS-CoV-2 has the potential to lead to post-COVID-19 condition, which is associated with reduced working capacity and absences from work.
- It is unclear if infection with SARS-CoV-2 leads to economic inactivity among those who were initially in employment just before the pandemic.

*What this study adds:*

- Infection is only very weakly or not associated with economic inactivity and non-employment in people who were in employment just prior to the pandemic.

*How this study might affect research, practice or policy:*

- Alternative explanations for the rise in economic inactivity need to be investigated such as long-term sickness.
- Possible biases introduced by linkage consent need considering when using linked healthcare data.

## Introduction

Considerable economic disruption has occurred internationally since the start of the pandemic (1), with many people leaving or losing their jobs (2). In the UK, coinciding with the spread of COVID-19, the number of economically inactive people increased dramatically, with 500,000 additional people being economically inactive when economic inactivity peaked in 2022 (3). This has been driven principally by people over the age of 50, with the number of older workers on unemployment-related benefits nearly doubling as a result of the pandemic (4), representing the largest increase in inactivity since records began in 1971 (5). At the macro-level, the UK’s labour market shortages have increased, while at the individual-level this may have had devastating financial and personal consequences for the people affected (6).

The health consequences of experiencing COVID-19 are possible drivers of increased economic inactivity. Post-COVID-19 condition (also known as ‘long COVID’) has been linked to reductions in working capacity (7), substantial absences from work (8–10), and disrupted finances (11). However, the studies in this area are generally limited, with most being based on small clinical samples consisting only of people who have been hospitalised for COVID-19 (12) or treated by primary care services (13), or cross-sectional surveys of people who have been recruited because they reported having had COVID-19 (8, 9). While there is a small number of other studies that have comparison groups, these studies also have limited ability to adjust for important sociodemographic factors (10, 14). Furthermore, while post-COVID-19 condition is the cause of problems that policy makers would like quantified, measures of it may not always be the best way of understanding the role of COVID-19 and subsequent employment outcomes. Post-COVID-19 condition is partly defined on the basis that symptoms impact on aspects of everyday functioning including work (15), thus post-COVID-19 condition could for some people be partly a consequence of their employment status, and other possible explanations for the rise in economic inactivity cannot be discounted (5). Investigating the impact of COVID-19 on economic activity is therefore challenging.

One approach is to use administrative data. Administrative data that combines healthcare data with data on labour market and other domains required to adjust for potentially sociodemographic confounding variables is not readily available (16). Longitudinal population surveys (LPS) are an important alternative source not only of economic activity data following the pandemic, but also socioeconomic conditions and health status prior to the pandemic, allowing for better adjustment of confounding. However, neither economic inactivity nor COVID-19 infection are typically common enough for analyses to be conducted using data from a single longitudinal study. A solution to these challenges is to pool data from multiple surveys and link them to NHS data to provide standardised measures for SARS-CoV-2 positivity across studies. The UK Longitudinal Linkage Collaboration (UK LLC) Trusted Research Environment (TRE), which has linked NHS data to many major UK longitudinal surveys (17), makes pooled analysis feasible.

Using data available in the UK LLC, we aim to understand the relationship that COVID-19 status as indicated by a positive test for SARS-CoV-2 has with economic inactivity and employment status, among respondents who reported that they were employed or self-employed just before the pandemic. The use of linked UK LLC data is not without restrictions that might bias the sample; data was limited for some LPSs due to lack of consent for linkage. To evaluate the possible consequences of potential consent bias, we carried out analyses using unlinked data from UK Data Service (UKDS) for the same LPS and used self-reported measures of COVID-19 instead of linked health records.

## Methods

We use self-reported sociodemographic, economic and COVID-19 data on individuals from five UK population-based longitudinal studies where the self-reported data are both held in the UK LLC (where they are linked to NHS health records) and the UKDS (where data from a wider sample, not limited by linkage consent, are held). These studies include three cohort studies (age-homogenous within-study): the 1970 British Cohort Study (BCS70) (18), the 1958 National Child Development Study (NCDS) (19) and Next Steps (NS, formerly known as the Longitudinal Study of Young People in England (20)). The studies also include two population surveys (age-heterogenous within-study): Understanding Society (USoc (21, 22)) and the English Longitudinal Study of Ageing (ELSA) (23). Details of the longitudinal studies are given in Table 1, with further descriptions in Appendix A of the Supplementary File. The UK LLC TRE hosts de-identified data and includes self-reported survey data linked to participants’ English data on positive tests for SARS-CoV-2 from the NHS Digital COVID-19 Second Generation Surveillance (COVIDSGSS), NHS Digital Npex, and NHS Digital Covid-19 UK Non-hospital Antibody Testing Results (17).

**Table 1:**
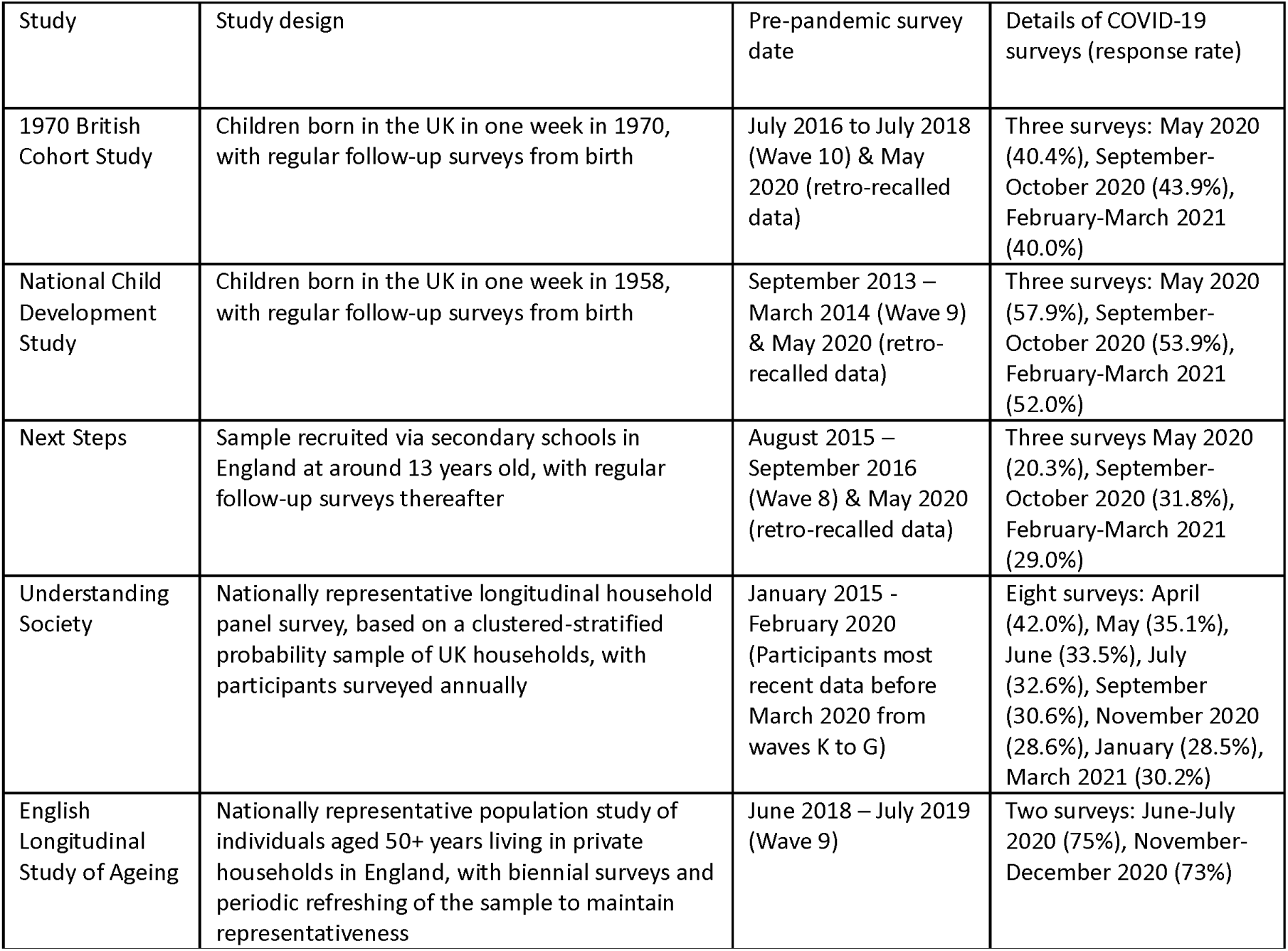
Survey dates for the longitudinal population studies used in the analyses.

| Study | Study design | Pre-pandemic survey date | Details of COVID-19 surveys (response rate) |
| --- | --- | --- | --- |
| 1970 British Cohort Study | Children born in the UK in one week in 1970, with regular follow-up surveys from birth | July 2016 to July 2018 (Wave 10) & May 2020 (retro-recalled data) | Three surveys: May 2020 (40.4%), September-October 2020 (43.9%), February-March 2021 (40.0%) |
| National Child Development Study | Children born in the UK in one week in 1958, with regular follow-up surveys from birth | September 2013 – March 2014 (Wave 9) & May 2020 (retro-recalled data) | Three surveys: May 2020 (57.9%), September-October 2020 (53.9%), February-March 2021 (52.0%) |
| Next Steps | Sample recruited via secondary schools in England at around 13 years old, with regular follow-up surveys thereafter | August 2015 – September 2016 (Wave 8) & May 2020 (retro-recalled data) | Three surveys May 2020 (20.3%), September-October 2020 (31.8%), February-March 2021 (29.0%) |
| Understanding Society | Nationally representative longitudinal household panel survey, based on a clustered-stratified probability sample of UK households, with participants surveyed annually | January 2015 - February 2020 (Participants most recent data before March 2020 from waves K to G) | Eight surveys: April (42.0%), May (35.1%), June (33.5%), July (32.6%), September (30.6%), November 2020 (28.6%), January (28.5%), March 2021 (30.2%) |
| English Longitudinal Study of Ageing | Nationally representative population study of individuals aged 50+ years living in private households in England, with biennial surveys and periodic refreshing of the sample to maintain representativeness | June 2018 – July 2019 (Wave 9) | Two surveys: June-July 2020 (75%), November-December 2020 (73%) |

Each longitudinal study sent surveys to their participants during the COVID-19 pandemic. These surveys contained demographic, health and COVID-19 related questions, and gathered information on employment status just before the pandemic, and employment status and economic activity during the pandemic. We derived two analytic samples using data pooled from the five longitudinal studies. Within the UK LLC TRE we selected people in England with permission to link to NHS data, aged 25 years and over in the first COVID-19 survey they participated in, and aged 65 or under at the time of assessing their economic activity and employment status at end of follow up (“UKLLC sample”). The younger age limit was selected to exclude those of younger ages when education would be a common alternative to employment. The older limit reflects statutory retirement age. In addition, we repeat analyses using a sample based on data available from the UKDS, from which the UKLLC sample was derived, without the restrictions of being resident in England and consenting to linkage to English NHS data (“UKDS sample”)

### Outcomes

The outcomes were economic inactivity (economically active -reference category - versus economically inactive) and employment status (in paid employment -reference category - versus not in employment). The distinction between the two measures is that being economically active is defined to include not only those who are in paid employment but also those who are unemployed and actively seeking work. In contrast for the employment status measure, the non-employed category includes both unemployed people who are looking for work and economically inactive people e.g. retired or long-term sick. To be as consistent as possible, we sought to use data from approximately the same period of each longitudinal cohort survey, which ranged from November/December 2020 to March 2021 (details are given in Table 1). More extensive details on how the outcome variables were defined are given in Appendix B in the Supplementary File.

### Exposure(s)

Two measures of COVID-19 status were used. For the UK LLC sample, the primary exposure was an NHS record indicating that participants tested positive for SARs-CoV-2 prior to the follow-up survey, during which economic inactivity or employment status was ascertained. For the UKDS sample, participants were categorised as not having COVID-19, having suspected COVID-19, or having test-confirmed COVID-19, according to self-reported measures up to and including the follow-up survey (see Appendix C).

### Potential confounding variables

We had available the following demographic variables: age at time of outcome (standardised linear and quadratic terms), sex (male versus female), household composition at start of pandemic (alone, partner no children, partner and children/grandchildren, children/grandchildren without partner, other (e.g. living with housemate(s)), self-reported key worker status during the pandemic (no/yes) and ethnicity (White, Asian, Black, Mixed, Other). NHS records, if available, were used if ethnicity data was missing (5.4% of the sample) for the UKLLC sample. For the UKDS analyses ethnicity was not used. BCS70 ethnicity data was restricted to participants of a sweep that occurred in the year 2000, and only a simple non-white binary measure was available for ELSA. Adjusting for ethnicity would have led to a smaller sample size for the addition of a variable of limited utility. The pre-pandemic socioeconomic confounders were the National Statistics Socio-Economic Classification (NS-SEC: higher management and professional, lower management and professional, intermediate, small employer, lower supervisory and technical, semi-routine, routine, unclassifiable) and the highest level of education (harmonised using National Vocational Qualifications (NVQ) or academic equivalents see Schneider 2011 (24) into NVQ4 or 5, NVQ3, NVQ2 or 1, none, unclassifiable). To adjust for health, we used pre-pandemic self-rated health (excellent, very good, good, fair or poor) and whether a person reported having been advised to ‘shield’ (i.e., self-isolate; no versus yes) during the pandemic, based on whether or not they reported receiving a letter from the NHS indicating that they were at risk of severe illness if they caught coronavirus. NS-SEC and self-rated health data for ELSA and USoc were collected prior to the pandemic, whereas retrospective measures collected during the first pandemic sweep were used for NCDS, BCS70 and NS.

### Statistical analyses

Analyses consisted of separate logistic regression models for the association between COVID-19 status and economic inactivity and employment status. Unadjusted and confounder adjusted analyses were conducted using the differing COVID-19 measures for the UKLLC and UKDS samples. Given that we pooled data from multiple studies with different study designs and did not have information on those who had not consented to linkage, we were not able to provide weighted estimations.

To investigate the comparability of the self-reported and NHS testing data, we conducted two sets of additional sensitivity analyses. First, using the UKLLC sample we repeated the analyses using self-reported COVID-19 status as the exposure. Second, we ran the analyses using only people resident in England in the UKDS sample. In addition, we carried out analyses stratifying the UKDS sample by the following characteristics: age (under 50 years versus 50 and over), sex and NS-SEC (higher management, administrators, and professionals versus intermediate, service and routine), and self-rated health (excellent, very good, good vs fair or poor). We formally tested for effect modification by these characteristics using likelihood ratio tests.

The code to derive all the variables and carry out the analyses is available from the UK LLC Github repository (https://github.com/UKLLC/llc_0010). The analyses and coding of the survey data were carried out using Stata 17.0, while the graphs and coding of NHS data were carried out using R 4.3.1

## Results

The UKLLC sample included 8,174 people who had complete data (full details on the selection process are given in Supplementary Figure 1a) of whom 479 people (5.9%) had an NHS record of a positive test for SARS-CoV-2, while 528 (6.5%) self-reported COVID-19 confirmed by a test and 1,164 (14.3%) suspected they had had COVID-19 (Table 2). The UKDS sample included 13,881 people (see Supplementary Figure 1b) for whom the percentages of people reporting COVID-19 confirmed by a test (6.4%, n= 881) or suspected COVID-19 (13.8% n = 1,918) were slightly lower than the UKLLC sample. The percentage of people reporting that they were economically inactive was similar in the UKLLC sample (5.3%, n= 429) and UKDS samples (5.0%, n = 689), as was the case for people reporting that they were not in work (UKLLC: 10.4%, n = 847; UKDS: 9.9%, n= 1,380). The majority of the UKLLC and UKDS samples were over the age of 50 (UKLLC: 71.5% n = 5,841, UKDS: 9,547, 68.8%). Both samples also over-represented females (UKLLC: 56.7% n = 4,6315, UKDS: 57.4%, n = 7,964).

**Table 2:**
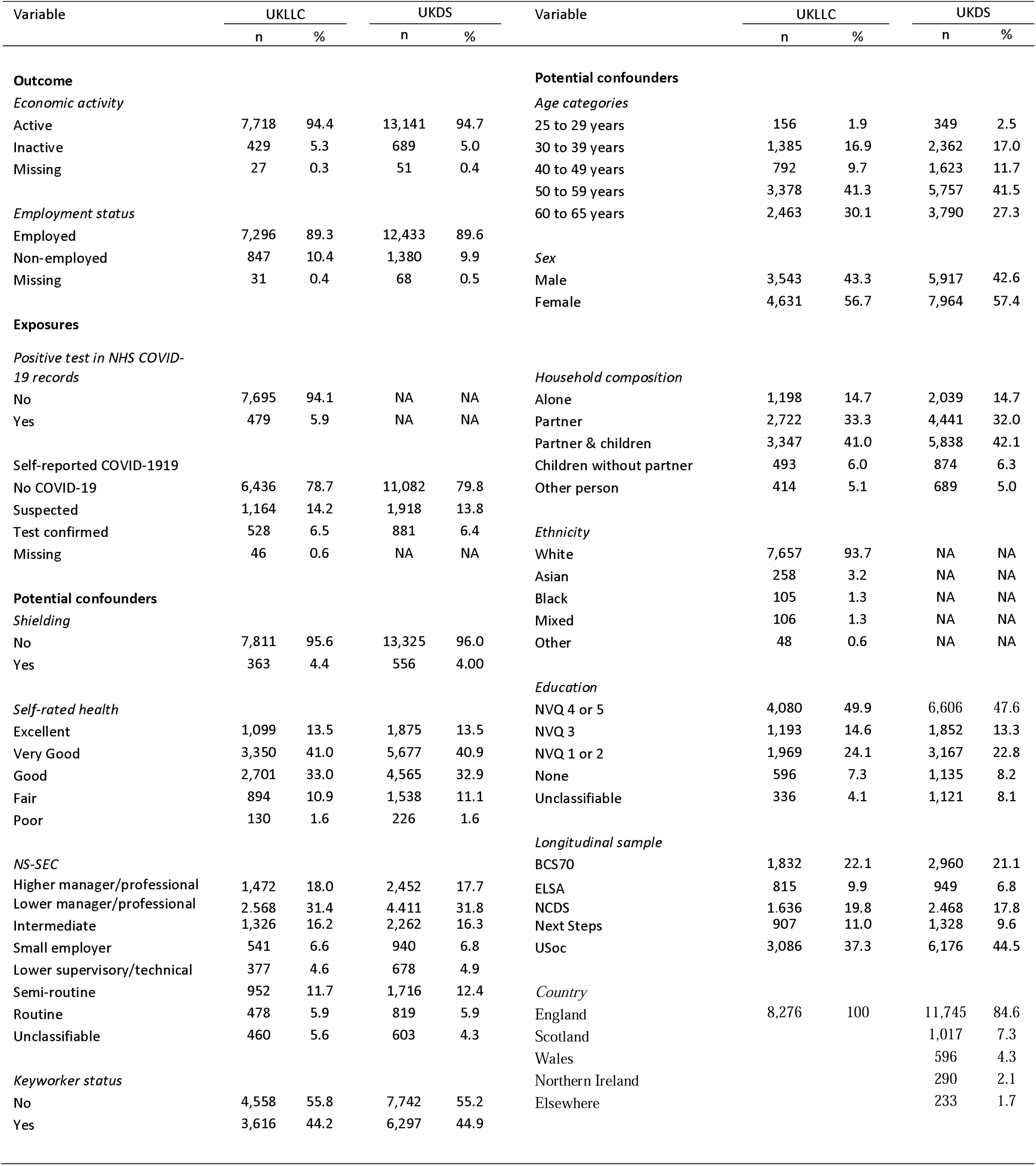
Characteristics of participants in the UKLLC and UKDS analytic samples.

| Variable | UKLLC |  | UKDS |  | Variable | UKLLC |  | UKDS |  |
| --- | --- | --- | --- | --- | --- | --- | --- | --- | --- |
|  | n | % | n | % |  | n | % | n | % |
| **Outcome** |  |  |  |  | **Potential confounders** |  |  |  |  |
| *Economic activity* |  |  |  |  | *Age categories* |  |  |  |  |
| Active | 7,718 | 94.4 | 13,141 | 94.7 | 25 to 29 years | 156 | 1.9 | 349 | 2.5 |
| Inactive | 429 | 5.3 | 689 | 5.0 | 30 to 39 years | 1,385 | 16.9 | 2,362 | 17.0 |
| Missing | 27 | 0.3 | 51 | 0.4 | 40 to 49 years | 792 | 9.7 | 1,623 | 11.7 |
|  |  |  |  |  | 50 to 59 years | 3,378 | 41.3 | 5,757 | 41.5 |
|  |  |  |  |  | 60 to 65 years | 2,463 | 30.1 | 3,790 | 27.3 |
| *Employment status* |  |  |  |  | *Sex* |  |  |  |  |
| Employed | 7,296 | 89.3 | 12,433 | 89.6 | Male | 3,543 | 43.3 | 5,917 | 42.6 |
| Non-employed | 847 | 10.4 | 1,380 | 9.9 | Female | 4,631 | 56.7 | 7,964 | 57.4 |
| Missing | 31 | 0.4 | 68 | 0.5 |  |  |  |  |  |
| **Exposures** |  |  |  |  |  |  |  |  |  |
| *Positive test in NHS COVID-19 records* |  |  |  |  | *Household composition* |  |  |  |  |
| No | 7,695 | 94.1 | NA | NA | Alone | 1,198 | 14.7 | 2,039 | 14.7 |
| Yes | 479 | 5.9 | NA | NA | Partner | 2,722 | 33.3 | 4,441 | 32.0 |
|  |  |  |  |  | Partner & children | 3,347 | 41.0 | 5,838 | 42.1 |
| Self-reported COVID-19 |  |  |  |  | Children without partner | 493 | 6.0 | 874 | 6.3 |
| No COVID-19 | 6,436 | 78.7 | 11,082 | 79.8 | Other person | 414 | 5.1 | 689 | 5.0 |
| Suspected | 1,164 | 14.2 | 1,918 | 13.8 |  |  |  |  |  |
| Test confirmed | 528 | 6.5 | 881 | 6.4 | *Ethnicity* |  |  |  |  |
| Missing | 46 | 0.6 | NA | NA | White | 7,657 | 93.7 | NA | NA |
|  |  |  |  |  | Asian | 258 | 3.2 | NA | NA |
| **Potential confounders** |  |  |  |  | Black | 105 | 1.3 | NA | NA |
| *Shielding* |  |  |  |  | Mixed | 106 | 1.3 | NA | NA |
| No | 7,811 | 95.6 | 13,325 | 96.0 | Other | 48 | 0.6 | NA | NA |
| Yes | 363 | 4.4 | 556 | 4.00 |  |  |  |  |  |
|  |  |  |  |  | *Education* |  |  |  |  |
| Self-rated health |  |  |  |  | NVQ 4 or 5 | 4,080 | 49.9 | 6,606 | 47.6 |
| Excellent | 1,099 | 13.5 | 1,875 | 13.5 | NVQ 3 | 1,193 | 14.6 | 1,852 | 13.3 |
| Very Good | 3,350 | 41.0 | 5,677 | 40.9 | NVQ 1 or 2 | 1,969 | 24.1 | 3,167 | 22.8 |
| Good | 2,701 | 33.0 | 4,565 | 32.9 | None | 596 | 7.3 | 1,135 | 8.2 |
| Fair | 894 | 10.9 | 1,538 | 11.1 | Unclassifiable | 336 | 4.1 | 1,121 | 8.1 |
| Poor | 130 | 1.6 | 226 | 1.6 |  |  |  |  |  |
|  |  |  |  |  | *Longitudinal sample* |  |  |  |  |
| NS-SEC |  |  |  |  | BCS 70 | 1,832 | 22.1 | 2,960 | 21.1 |
| Higher manager/professional | 1,472 | 18.0 | 2,452 | 17.7 | ELSA | 815 | 9.9 | 949 | 6.8 |
| Lower manager/professional | 2,568 | 31.4 | 4,411 | 31.8 | NCDS | 1,636 | 19.8 | 2,468 | 17.8 |
| Intermediate | 1,326 | 16.2 | 2,262 | 16.3 | Next Steps | 907 | 11.0 | 1,328 | 9.6 |
| Small employer | 541 | 6.6 | 940 | 6.8 | USoc | 3,086 | 37.3 | 6,176 | 44.5 |
| Lower supervisory/technical | 377 | 4.6 | 678 | 4.9 |  |  |  |  |  |
| Semi-routine | 952 | 11.7 | 1,716 | 12.4 | *Country* |  |  |  |  |
| Routine | 478 | 5.9 | 819 | 5.9 | England | 8,276 | 100 | 11,745 | 84.6 |
| Unclassifiable | 460 | 5.6 | 603 | 4.3 | Scotland |  |  | 1,017 | 7.3 |
|  |  |  |  |  | Wales |  |  | 596 | 4.3 |
| Keyworker status |  |  |  |  | Northern Ireland |  |  | 290 | 2.1 |
| No | 4,558 | 55.8 | 7,742 | 55.2 | Elsewhere |  |  | 233 | 1.7 |
| Yes | 3,616 | 44.2 | 6,297 | 44.9 |  |  |  |  |  |

### Economic inactivity

In the UKLLC sample having an NHS record of a positive test for SARS-CoV2 was not significantly associated with reduced risk economic inactivity in unadjusted analyses (odds ratio (OR) 0.82, 95% confidence interval (CI) 0.52 to 1.28) and there was a weak non-significant increased risk in adjusted analyses (OR 1.08, 95% CI 0.68 to 1.73) (see the red circle in Panel A of Figure 1 and Supplementary Table 1).

**Figure 1:**
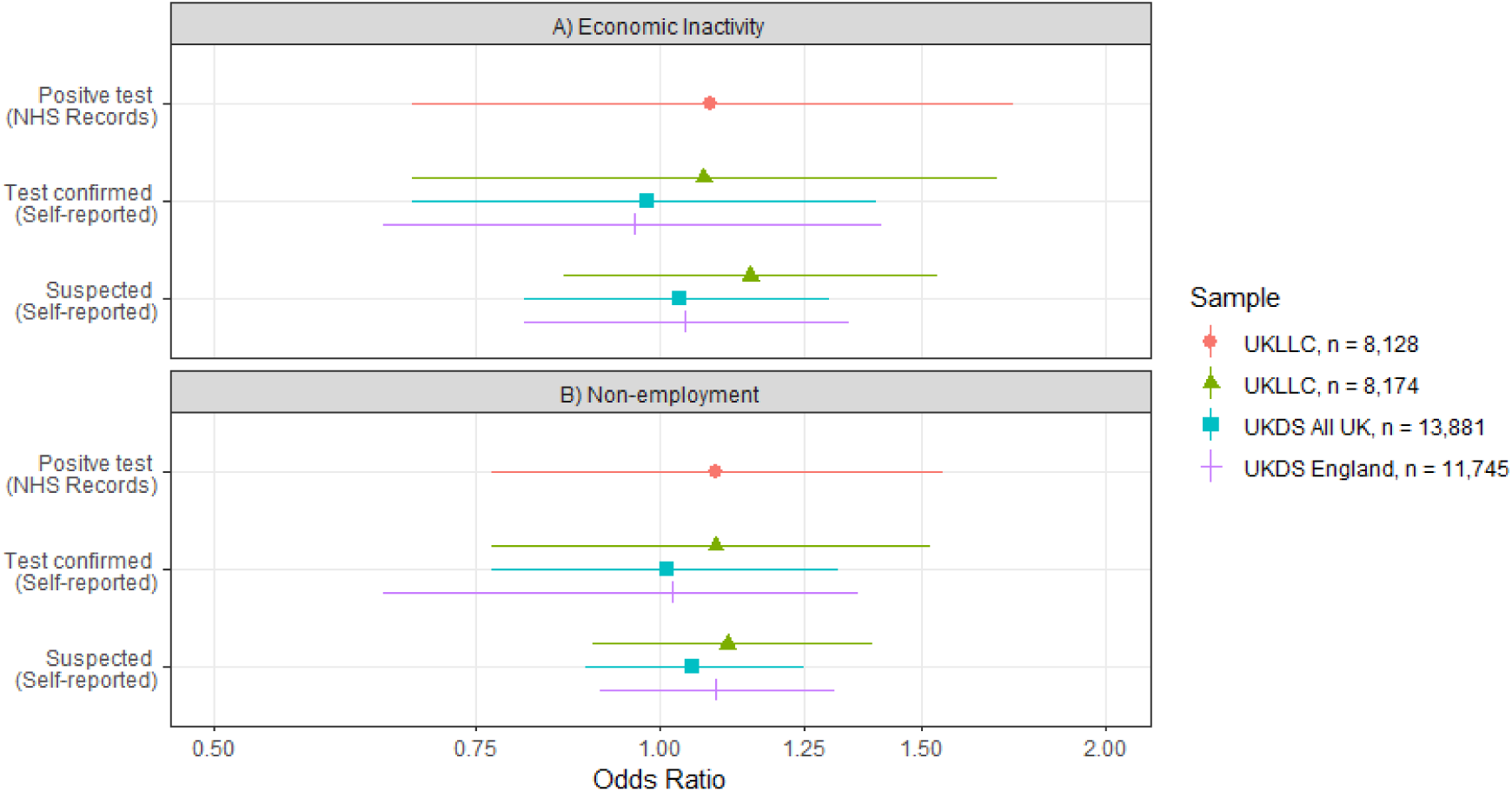
Odds ratios for economic inactivity and non-employment by COVID-19 status in 4 subsamples of 5 UK Longitudinal Population Studies. Analyses were adjusted for age, sex, household composition, ethnicity (UKLLC only), longitudinal study, country (UKDS only), shielding status, self-rated health, NS-SEC, keyworker status and education.

In the UKDS sample self-reported test confirmed COVID-19 had a non-significant protective association with economic inactivity in unadjusted analyses (OR 0.77, 95% CI 0.54 to 1.09) that became null on adjustment (OR 1.01, 95% CI 0.70 to 1.44) analyses. Likewise, there was little evidence that suspected COVID-19 was associated with economic inactivity in unadjusted (OR 0.87 95% CI 0.69 to 1.10) or adjusted analyses (OR 1.02 95% 0.80 to 1.29) (see the blue squares in Panel A of Figure 1 and Supplementary Table 2).

### Employment status

In the UKLLC sample a positive test record for SARS-CoV-2 was associated with an increased risk of not being in employment in unadjusted (OR 0.77, 95% CI 0.56 to 1.08) and only a very week non-significant increased risk in adjusted analyses (OR 1.09, 95% CI 0.77 to 1.55) (see the red circle in Figure 1 Panel B, and Supplementary Table 1).

In the UKDS sample, self-report of test confirmed COVID-19 was associated with reduced chance of being without a job status in unadjusted analyses (OR 0.72, 95% CI 0.56 to 0.93) but there was little evidence of an association in adjusted analyses (OR 1.03, 95% 0.79 to 1.35). There was little evidence that suspected COVID-19 was associated with employment status in unadjusted (OR 0.89, 95% 0.76 to 1.06) and a week non-significant association in adjusted estimates (OR 1.07, 95% 0.90 to 1.28) (see blue squares in Figure 1 Panel B, and Supplementary Table 2).

### Additional sensitivity analyses

The adjusted associations for both economic inactivity and employment status for different samples and different COVID-19 status measure combinations are also shown in Figure 1. Given the moderately wide confidence intervals, associations in either direction cannot be ruled out completely. The results for positive test for SARs-CoV-2 as indicated by test records (red circles) and self-reported test-confirmed COVID-19 in the UKLLC sample (green triangles) show very similar relationships of a very small increased risk of economic inactivity or non-employment. The self-reported test-confirmed measure shows a similar null association for the UKDS sample included the whole of the UK (blue square) and restricted to England only (violet crosses). Albeit with similar cautions as the confidence intervals were only slightly narrower.

The relationship between COVID-19 status and the economic inactivity and employment status measures stratified by age, sex, NS-SEC and self-reported health are shown in Table 3. Overall, neither the stratified results nor formal testing supported the idea that the relationship between COVID-19 and economic inactivity or employment status varied by any of these factors.

**Table 3:** Adjusted^1^ associations between self-reported COVID-19 status and economic inactivity and non-employment stratified by age, sex, NS-SEC, self-reported health and keyworker status using the UKDS full sample

| COVID-19 status | Economic Inactivity |  |  |  | Non-employment |  |  |  |
| --- | --- | --- | --- | --- | --- | --- | --- | --- |
| Ref = No Covid | Odds Ratio | 95% CI |  | p | Odds Ratio | 95% CI |  | p |
| Age |  |  |  |  |  |  |  |  |
| People under 50 |  |  |  |  |  |  |  |  |
| Suspected | 0.93 | 0.53 | 1.63 | 0.794 | 1.23 | 0.86 | 1.76 | 0.260 |
| Test confirmed | 1.35 | 0.68 | 2.67 | 0.393 | 1.07 | 0.62 | 1.85 | 0.801 |
| People 50 and over |  |  |  |  |  |  |  |  |
| Suspected | 1.03 | 0.79 | 1.34 | 0.835 | 1.03 | 0.84 | 1.25 | 0.804 |
| Test confirmed | 0.92 | 0.60 | 1.40 | 0.699 | 1.03 | 0.75 | 1.40 | 0.855 |
| Sex |  |  |  |  |  |  |  |  |
| Male |  |  |  |  |  |  |  |  |
| Suspected | 1.06 | 0.71 | 1.56 | 0.789 | 1.11 | 0.84 | 1.46 | 0.471 |
| Test confirmed | 0.91 | 0.48 | 1.72 | 0.765 | 1.16 | 0.46 | 1.23 | 0.262 |
| Female |  |  |  |  |  |  |  |  |
| Suspected | 1.01 | 0.74 | 1.36 | 0.971 | 1.06 | 0.84 | 1.33 | 0.642 |
| Test confirmed | 1.05 | 0.68 | 1.62 | 0.830 | 1.21 | 0.87 | 1.68 | 0.257 |
| NS-SEC |  |  |  |  |  |  |  |  |
| Higher occupations |  |  |  |  |  |  |  |  |
| Suspected | 1.28 | 0.92 | 1.79 | 0.144 | 1.33 | 1.03 | 1.71 | 0.003 |
| Test confirmed | 1.14 | 0.67 | 1.95 | 0.629 | 0.99 | 0.65 | 1.55 | 0.988 |
| Intermediate and routine occupations |  |  |  |  |  |  |  |  |
| Suspected | 0.80 | 0.55 | 1.17 | 0.250 | 0.95 | 0.74 | 1.22 | 0.687 |
| Test confirmed | 0.97 | 0.60 | 1.62 | 0.940 | 0.96 | 0.69 | 1.42 | 0.957 |
| Self-rated health |  |  |  |  |  |  |  |  |
| Excellent or good health |  |  |  |  |  |  |  |  |
| Suspected | 1.10 | 0.84 | 1.42 | 0.492 | 1.11 | 0.92 | 1.34 | 0.281 |
| Test confirmed | 0.87 | 0.57 | 1.32 | 0.513 | 0.94 | 0.71 | 1.31 | 0.820 |
| Fair or poor health |  |  |  |  |  |  |  |  |
| Suspected | 0.76 | 0.41 | 1.40 | 0.373 | 0.97 | 0.65 | 1.49 | 0.899 |
| Test confirmed | 1.67 | 0.81 | 3.44 | 0.167 | 1.43 | 0.77 | 2.64 | 0.255 |
| Key worker status |  |  |  |  |  |  |  |  |
| Not keyworkers |  |  |  |  |  |  |  |  |
| Suspected | 1.10 | 0.83 | 1.45 | 0.504 | 1.14 | 0.93 | 1.39 | 0.200 |
| Test confirmed | 1.16 | 0.78 | 1.85 | 0.409 | 1.09 | 0.78 | 1.51 | 0.611 |
| Key workers |  |  |  |  |  |  |  |  |
| Suspected | 0.83 | 0.51 | 1.34 | 0.440 | 0.91 | 0.62 | 1.34 | 0.632 |
| Confirmed | 0.72 | 0.37 | 1.39 | 0.329 | 0.96 | 0.59 | 1.56 | 0.868 |

<sup>1.</sup> Analyses were adjusted for age, sex, household composition, longitudinal study, country, shielding status, self-rated health, NS-SEC, keyworker status, and education (stratification variable was omitted).

## Discussion

Our results come from five UK longitudinal population studies covering people in work at the start of the pandemic and adjusting for a wide range of potential confounders. They suggest that having SARs-CoV-2 confirmed by a test is either not associated or only has a small increased risk of being economically inactive and not being in work during the pandemic. There was also little evidence to suggest that this varied by important characteristics such as age, sex, and gender. Despite the reasonably large sample sizes for the samples, approximately 8,000 for the UKLLC sample and 14,000 for the UKDS sample, the width of confidence intervals does not rule out stronger associations entirely.

The finding, for people employed prior to the pandemic, that COVID-19 has a small or null association with economic inactivity and non-employment is not entirely inconsistent with the current literature, which is in its early stages of development, and currently focused on post-COVID condition. A non-peer reviewed initial rapid scoping review suggests that the socioeconomic consequences of post COVID-19 are considerable (12), however, the review also indicated that long-term absences from work may only occur for 20% of people with post COVID-19 condition. Post-COVID-19 condition itself only arises in a small proportion of those COVID-19 cases (25), and any possible impact is due to large numbers of people being infected with COVID-19 (12). Studies such as Ayoubkhani et al. (10) that observed a relationship between post COVID-19 condition and economic inactivity, have included both those who were employed and not in work prior to the pandemic. It is possible that the consequences of COVID-19 may be greater for those on the periphery of the labour market. The principal initial drivers of the rise in economic inactivity appear to be age and poor health (3), which we included as potential confounders and did find associations consistent with this. Explanations for difference between the UKLLC sample and the UKDS sample merit discussion. The difference is unlikely to be the result of using different indicators for COVID-19 status. For the UKLLC sample, the results using both the self-reported and NHS records are almost the same. Similarly, the differences between the UKLS and UKDS are unlikely to be due to the geographic differences between the samples. There was little difference between the associations for the self-reported measure of COVID-19 between the UKDS sample restricted to England and the sample for the whole of the UK. One possible explanation is the requirement for consent to NHS linkage.

For some of the studies included in the UK LLC, linkage to NHS records was based on consent. For most studies, including BCS70, NCDS, ELSA, and NS, consent for record linkage was collected prior to the COVID-19 pandemic. USoc local governance requirements demanded specific consent for inclusion in UK LLC, and this was acquired during the 8^th^ pandemic survey in March 2021. It is possible that differences between those who consented to linkage and those who did not may have introduced a small bias, which would be consistent with other studies (26). Given that data in the UK LLC is only available for those who consented, it is not currently possible to model those biases directly. However, there are plans to obtain consent for more people. Given the current data limitations, we believe that the sensitivity analysis comparing the UKDA sample and UKLLC sample is a reasonable alternative.

### Strengths and limitations

The use of a directly observed positive test for SARS-CoV-2 and a self-reported measure of test-confirmed COVID-19 each come with advantages and disadvantages. Conceptually, test-confirmed COVID-19 is much more common than other potential measures related to COVID-19 such as post COVID-19 condition, which is experienced by only small proportion of those who are infected (25). In addition, test-confirmed SARS-CoV-2 is not dependent on criteria related to daily functioning which may include employment (15), which are likely to be the result of multiple causes not just COVID-19.

However, the use of a test to ascertain COVID-19 does mean that poorer access to testing is a possible source of bias that could lead to an underestimation of effects (27); we hope adjusting for keyworker status minimised this. The use of test-confirmed SARS-CoV-2 will include asymptomatic, mild cases of COVID-19, and may exclude those unable or unwilling to be tested, thus would be expected to have weaker associations than measures of more severe COVID-19. A second weakness is that we only have information on economic inactivity and employment status covering the early pandemic periods, until March 2021. This period was before most people had access to vaccines and where COVID-19 posed the greatest health risk. However, it was also a period during which other policies and practices were in place such as furlough (28) and homeworking (29), and these may have enabled people to stay economically active.

Surveys are good at capturing snapshots around the time the survey measure is recorded. A strength of our study is that the use of longitudinal survey data meant we could control for important pre-pandemic potential confounding factors such as education and occupational class. However, it is possible that we could not fully adjust for factors that might lead to behavioural changes reducing the risk of exposure to the SARs-CoV-2 virus and labour market engagement. To mitigate against this, we did adjust for shielding, keyworker status, age and self-rated health. The UK LLC is planning to add linked administrative employment data which may extend the scope and period of employment outcomes that can be investigated. However, use of administrative data is not without other sources of bias (16).

For our chosen LPS, the UKLLC sample only included people who had consented to having their survey data linked to NHS records. In addition, despite being embedded within long standing cohorts, survey responses during the pandemic were lower than typically achieved. Thus, there may be some selection biases restricting generalisability of results and we did find slightly higher point estimates for the associations between suspected COVID-19 and the outcomes in the UKLLC sample, than the more general population sample. Given that we only have data within the UK LLC on people who had consented to linkage, options were limited. Regression analyses tend to be robust to missing data and more appropriate than other alternatives such as weighting and multiple imputation which could amplify biases when the data is missing not at random (30, 31). There is also the possibility that linkage errors and other errors with administrative data could potentially result in misclassification bias (16). It should be noted that for the UKLLC sample, slightly more people (see Supplementary Table 3) reported having had COVID-19 confirmed with a test in the self-reported data, than in the NHS test records.

The study also has many strengths. We have focused on people aged over 25 and in employment just prior to the pandemic. Thus, we are looking at the consequences of testing positive for COVID-19 in the group whose subsequent employment status and inactivity are mostly like to be due to the consequences of the pandemic. Data include pre-pandemic measures of NS-SEC and self-rated health. While the pre-pandemic measures are assessed in the first COVID-19 wave for NCDS, BCS70 and NS, this is still prior to the assessment of the employment outcome and thus longitudinal.

Finally, pooling data across studies has enabled a much larger sample size than any of the individual studies alone would have allowed. Using the UK LLC, pooling data has enabled the use of more detailed ethnicity information than is commonly available for BCS70, ELSA and NCDS, and a more detailed level of education and social class than employed in other approaches such as pooled meta-analysis, which have been used to combine longitudinal population studies in other papers (29).

### Interpretation and conclusions

Our results suggest either a small or no effect of COVID-19 among people who were employed during the pandemic. The direct consequences of SARS-CoV-2 infection are unlikely to be a major contributor to the large increases in economic inactivity seen following the pandemic, and therefore other explanations need to be investigated. Given the potentially small impact of COVID-19 among the working population, definitive answers are likely to require very large studies. This may require investments in studies to evaluate population health on a scale that was inconceivable prior to the pandemic. Alternatively, research should specifically target more vulnerable groups such as older people, or those already in poor health.

## Data Availability

Data used in this research are made available via UK Longitudinal Linkage Collaboration (UK LLC), which is a Trusted Research Environment developed and operated by the Universities of Bristol and Edinburgh using an underlying 'Secure eResearch Platform' provided by Swansea University for the processing and analysis of longitudinal study data with linked routine records. These data cannot be used or shared outside this environment. Researchers can apply to use UK LLC's resource using the procedure outlined in the UK LLC Data Access and Acceptable Use Policy (https://ukllc.ac.uk/governance/). The UK LLC uses a system of managed open access for researchers who demonstrate their project is intended to improve the public good.
For the survey data, anonymised datasets with corresponding documentation can be de downloaded from the UK Data Service. More information for each longitudinal population study is available in the Supplementary File in Appendix A.

https://ukllc.ac.uk/data-use-register

https://ukdataservice.ac.uk/

## Declarations

### Ethics approval

This project has been approved by UK LLC and its contributing data owners and information on this project and its outputs can be accessed via UK LLC’s website (Data Use Register | UK Longitudinal Linkage Collaboration; https://ukllc.ac.uk/data-use-register) and UK LLC’s GitHub (UK Longitudinal Linkage Collaboration GitHub; https://github.com/UKLLC). The UK LLC has ethical approval from the Health Research Authority Research Ethics Committee (Haydock Committee; ref: 20/NW/0446). More information on ethical approval for each longitudinal population study is available in the Supplementary File in Appendix A.

### Data sharing statement

Data used in this research are made available via UK Longitudinal Linkage Collaboration (UK LLC), which is a Trusted Research Environment developed and operated by the Universities of Bristol and Edinburgh using an underlying ‘Secure eResearch Platform’ provided by Swansea University for the processing and analysis of longitudinal study data with linked routine records. These data cannot be used or shared outside this environment. Researchers can apply to use UK LLC’s resource using the procedure outlined in the UK LLC Data Access and Acceptable Use Policy (https://ukllc.ac.uk/governance/) . The UK LLC uses a system of managed open access for researchers who demonstrate their project is intended to improve the public good.

For the survey data, anonymised datasets with corresponding documentation can be de downloaded from the UK Data Service. More information for each longitudinal population study is available in the Supplementary File in Appendix A.

### Authors’ contributions

RJSh, RR, RJSi, JW, JZ, OKLH, RCEB, BM, MJG, ED, FG, GBP, SVK were responsible for the conception and design. RJSh, RR, JZ, GDG, MJG, RJSi, AB, SVK were responsible for data curation and analyses. RJSh, SP created the graphs and figures. RJSh, RR, RJSi, JW, OKLH, AB, SVK, wrote the original draft of the manuscript. All authors were involved in revising the manuscript.

## Acknowledgements

The UK Longitudinal Linkage Collaboration (UK LLC) is a Trusted Research Environment (TRE) developed and operated by the Universities of Bristol and Edinburgh using an underlying ‘Secure eResearch Platform’ infrastructure (SeRP UK: https://serp.ac.uk/) provided by Swansea University for longitudinal research. The UK LLC TRE is designed to host de-identified data from many interdisciplinary, longitudinal population studies (LPS); to systematically link these to participants’ health, administrative and environmental records; and to provide a secure analysis environment. We thank the SeRP UK Team at Swansea University and NHS Digital Health and Care Wales for providing the TRE’s infrastructure and support.

This work uses data provided by participants of the contributing LPS within the UK LLC TRE, which have been collected through their longitudinal study or as part of their care and support and/or interactions with UK government services. We wish to recognise and thank the study participants and each contributing LPS team, including data managers, administrators and those collecting data. We thank the following LPS for contributing data that made this research possible: 1970 British Cohort Study (BCS70), English Longitudinal Study of Ageing (ELSA), National Child Development Study (NCDS), Next Steps, and the UK Household Longitudinal Study (Understanding Society).

We thank the NHS and particularly NHS Digital for their work in curating participants’ health records and for making these available for public benefit research designed to improve health services.’

A full list of acknowledgments, including support for each study, is provided in the Supplementary File in Appendix A.

