## Supplementary File for "Associations between SARS-CoV-2 infection and subsequent economic inactivity and employment status: pooled analyses of five linked longitudinal surveys"

### Supplementary Materials

### Appendix A: Ethics and data access statements for each study

#### BCS70: 1970 British Cohort Study

| **Description of Study Population** (including citations and references if required) | The 1970 British Cohort Study (BCS70) follows the lives of more than 17,000 people born in England, Scotland and Wales in a single week of 1970. Over the course of cohort members’ lives, BCS70 has collected information on health, physical, educational and social development, and economic circumstances, among other factors.  Since the birth survey in 1970, there have been nine ‘sweeps’ of all cohort members at ages 5, 10, 16, 26, 30, 34, 38, 42 and most recently at 46 (a biomedical data collection). The Age 51 Sweep is currently in the field (2022).  Data have been collected from a number of different sources, including the midwife present at birth, parents of the cohort members, head and class teachers, school health service personnel and the cohort members themselves.  The data have been collected in a variety of ways, including via paper and electronic questionnaires, clinical records, medical examinations, biological samples, physical measurements, tests of ability, educational assessments and diaries.  The study is conducted by the Centre for Longitudinal Studies. |
| --- | --- |
| **Acknowledgements** | BCS70 is core-funded by the ESRC. |
| **Ethics** | Ethics approval has been obtained for each follow-up from an NHS Research Ethics Committee (REC) since 2000. In addition, separate REC approval is in place to cover the ongoing activities of the study in between major sweeps of data collection (i.e. keeping in touch with and tracing cohort members; cleaning, documenting and providing access to the data for research; and linking data from administrative sources to survey data to increase the utility of the data for research). |
| **Website for Data Requests** | <https://cls.ucl.ac.uk/cls-studies/bcs70/> |

#### ELSA: English Longitudinal Study of Ageing

| **Description of Study Population** (including citations and references if required) | The English Longitudinal Study of Ageing (ELSA) is a unique and rich resource of information on the dynamics of health, social, wellbeing and economic circumstances in the English population aged 50 years and older ^1^.  The original sample was drawn from households that had previously responded to the Health Survey for England (HSE) between 1998 and 2001. The main fieldwork began in March 2002. The same group of respondents have been interviewed at two-yearly interviews.  ^1^Banks J, Batty GD, Breedvelt JJF, Coughlin K, Crawford R, Marmot M, Nazroo J, Oldfield Z, Steel N, Steptoe A, Wood M, Zaninotto P (2021) English Longitudinal Study of Ageing: Waves 0-9, 1998-2019 |
| --- | --- |
| **Acknowledgements** | The English Longitudinal Study of Ageing was developed by a team of researchers based at University College London, NatCen Social Research, the Institute for Fiscal Studies, the University of Manchester and the University of East Anglia. The data were collected by NatCen Social Research. The funding is currently provided by the National Institute on Aging (Ref: R01AG017644) and by a consortium of UK government departments, including Department for Health and Social Care; Department for Transport; and Department for Work and Pensions, which is coordinated by the National Institute for Health Research (NIHR, Ref: 198-1074). Funding has also been provided by the Economic and Social Research Council (ESRC). |
| **Ethics** | Ethical approval for the COVID-19 subsample was obtained from the University College London Research Ethics Committee (0017/003).  Ethical approval was received from the South Central -Berkshire Research Ethics Committee for Waves 9 (17/SC/0030) and 8 (15/SC/0526).  See also <https://www.elsa-project.ac.uk/ethical-approval> |
| **Website for Data Requests** | <https://www.elsa-project.ac.uk/data-and-documentation> |

#### NCDS: National Child Development Study

| **Description of Study Population** (including citations and references if required) | The National Child Development Study (NCDS) is a continuing longitudinal study that seeks to follow the lives of all those living in Great Britain who were born in one particular week in 1958. Conducted by the Centre for Longitudinal Studies, the aim of the study is to improve understanding of the factors affecting human development over the whole lifespan. It collects information on physical and educational development, economic circumstances, employment, family life, health behaviour, wellbeing, social participation and attitudes.  The broad aim of the study is to examine the impact that circumstances and experiences at one stage of life have on outcomes and achievements in later life. Since the birth survey in 1958, there have been ten ‘sweeps’ of all cohort members at ages 7, 11, 16, 23, 33, 42, 44/5 (a biomedical collection) 46, 50 and most recently at 55. The Age 62 Sweep is currently in the field (2022).  Data have been collected from a number of different sources, including the midwife present at birth, parents of the cohort members, teachers, doctors and the cohort members themselves. The data have been collected in a variety of ways, including via paper and electronic questionnaires, clinical records, medical examinations, biological samples, physical measurements, tests of ability and educational assessments.  The information collected forms a high quality data resource for scientific investigations across a full range of domains of individuals’ lives and across different points in time. The study has been designed to ensure comparability with other major cohort studies and to permit the examination of links between social change and the changing experiences of different cohorts.  <https://cls.ucl.ac.uk/cls-studies/1958-national-child-development-study/> |
| --- | --- |
| **Acknowledgements** | NCDS is core-funded by the ESRC. |
| **Ethics** | Ethics approval has been obtained for each follow-up from an NHS Research Ethics Committee (REC) since 2000. In addition, separate REC approval is in place to cover the ongoing activities of the study in between major sweeps of data collection (i.e. keeping in touch with and tracing cohort members; cleaning, documenting and providing access to the data for research; and linking data from administrative sources to survey data to increase the utility of the data for research). |
| **Website for Data Requests** | <https://cls.ucl.ac.uk/cls-studies/ncds/> |

#### NS: Next Steps

| **Description of Study Population** (including citations and references if required) | Next Steps (previously known as the Longitudinal Study of Young People in England (LSYPE1)) is a major longitudinal study that follows the lives of around 16,000 people born in 1989-90. The first seven sweeps of the study (2004-2010) were funded and managed by the Department for Education and mainly focused on the educational and early labour market experiences of young people.  The study began in 2004 and included young people in Year 9 who attended state and independent schools in England. Following the initial survey at age 13-14, the cohort members were interviewed every year until 2010.  In 2013 the management of Next Steps was transferred to the Centre for Longitudinal Studies (CLS) at the IOE, UCL’s Faculty of Education and Society. The first sweep conducted by CLS aimed to find out how the lives of the cohort members had turned out at age 25. It maintained the strong focus on education, but the content was broadened to become a more multi-disciplinary research resource.  The Age 32 Sweep is currently in the field (2022).  [https://doc.ukdataservice.ac.uk/doc/5545/mrdoc/pdf/next_steps_userguide_to_the_redeposit_of_sweeps_1to7_may2020.pdf](https://eur01.safelinks.protection.outlook.com/?url=https%3A%2F%2Fdoc.ukdataservice.ac.uk%2Fdoc%2F5545%2Fmrdoc%2Fpdf%2Fnext_steps_userguide_to_the_redeposit_of_sweeps_1to7_may2020.pdf&data=05%7C01%7Cmorag.henderson%40ucl.ac.uk%7Ca5279433ef414feb792808da6319282f%7C1faf88fea9984c5b93c9210a11d9a5c2%7C0%7C0%7C637931256931551677%7CUnknown%7CTWFpbGZsb3d8eyJWIjoiMC4wLjAwMDAiLCJQIjoiV2luMzIiLCJBTiI6Ik1haWwiLCJXVCI6Mn0%3D%7C3000%7C%7C%7C&sdata=ColBf1sx2sz8g95l4WKpe3z%2B1ymQyhVgJOvHoubF3BY%3D&reserved=0)  [https://doc.ukdataservice.ac.uk/doc/5545/mrdoc/pdf/nextsteps_age25_survey_user_guide_v3.pdf](https://eur01.safelinks.protection.outlook.com/?url=https%3A%2F%2Fdoc.ukdataservice.ac.uk%2Fdoc%2F5545%2Fmrdoc%2Fpdf%2Fnextsteps_age25_survey_user_guide_v3.pdf&data=05%7C01%7Cmorag.henderson%40ucl.ac.uk%7Ca5279433ef414feb792808da6319282f%7C1faf88fea9984c5b93c9210a11d9a5c2%7C0%7C0%7C637931256931551677%7CUnknown%7CTWFpbGZsb3d8eyJWIjoiMC4wLjAwMDAiLCJQIjoiV2luMzIiLCJBTiI6Ik1haWwiLCJXVCI6Mn0%3D%7C3000%7C%7C%7C&sdata=Xy8K4SuQRsqAMbqs19kgBiff4EQvnlHm6HANwXQY%2B20%3D&reserved=0)  <https://cls.ucl.ac.uk/cls-studies/next-steps/> |
| --- | --- |
| **Acknowledgements** | Next Steps now is core-funded by the ESRC. |
| **Ethics** | Ethics approval is obtained for each follow-up from an NHS Research Ethics Committee (REC). In addition, separate REC approval is in place to cover the ongoing activities of the study in between major sweeps of data collection (i.e. keeping in touch with and tracing cohort members; cleaning, documenting and providing access to the data for research; and linking data from administrative sources to survey data to increase the utility of the data for research). |
| **Website for Data Requests** | <https://cls.ucl.ac.uk/cls-studies/next-steps/> |

#### Understanding Society – the UK Household Longitudinal Study

| **Description of Study Population** (including citations and references if required) | Understanding Society, the UK Household Longitudinal Study, is a longitudinal survey of the members of ~40,000 households (at Wave 1, 2009-10) in the United Kingdom. The survey sample consists of a large General Population Sample (~26,000 households) plus three other components: the Ethnic Minority Boost Sample (~4,000 households), the former British Household Panel Survey sample (~8,000 households) and the Immigrant and Ethnic Minority Boost Sample (~2,900 households, added at Wave 6). Household and individual interviews are conducted annually. The study is multi-topic and multi-purpose.  From April 2020 to September 2021, participants from the main Understanding Society sample were asked to complete nine short web-surveys (with a telephone option in some months). The COVID-19 study covered the changing impact of the pandemic on the welfare of UK individuals, families and wider communities. ~18,000 individuals provided a full or partial interview at Wave 1 (April 2020).  At Wave 8 of the COVID-19 study, 8,477 participants provided consent to link their survey data to administrative health records. |
| --- | --- |
| **Acknowledgements** | Understanding Society is an initiative funded by the Economic and Social Research Council and various Government Departments, with scientific leadership by the Institute for Social and Economic Research, University of Essex, and survey delivery by NatCen Social Research and Kantar Public.  The COVID-19 study (2020-2021) was funded by the Economic and Social Research Council and the Health Foundation. Serology testing was funded by the COVID-19 Longitudinal Health and Wealth – National Core Study. Fieldwork for the web survey was carried out by Ipsos MORI and for the telephone survey by Kantar Public. |
| **Ethics** | The University of Essex Ethics Committee has approved all data collection on Understanding Society main study, COVID-19 surveys and innovation panel waves, including asking consent for all data linkages except for health records.  Approval for asking consent for health record linkage and for the collection of blood and subsequent serology testing in the March 2021 wave of the COVID-19 study was obtained from London – City & East Research Ethics Committee (21/HRA/0644). |
| **Website for Data Requests** | <https://ukllc.ac.uk> or <https://ukdataservice.ac.uk> |

### Appendix B: Coding of economic activity and employment status by each cohort

Participants in all cohorts were asked about their current employment status, and whether they were actively seeking jobs in two Cohorts (USoc and ELSA) – details are given below. *Economic activity* was coded as a binary variable (economically active - reference category - versus economically inactive), using both questions where available, and making assumptions about the employment status variables for each cohort. *Employment status* (in employment – reference category - versus not in employment) was coded using different assumptions. How each cohort is coded is described below.

#### CLS studies: BCS70, NCDS, NS

Participants were asked a single question: “We/I] would like to ask you about what you are currently doing. Which of [these/the following] would you say best describes your situation now?” Participants were classified into “Economic activity” and “Employment status” as follows:

##### Economic activity

Economically active (0: Employed and currently working (or on annual leave/holiday), Employed but on paid leave (including furlough), Employed and on unpaid leave, Apprenticeship, Self-employed and currently working (or on holiday) Unemployed).

Economically inactive (1: In unpaid/voluntary work, Permanently sick or disabled, Looking after home or family, In education at school/college/university, Retired, Doing something else).

##### Employment status

Employed (0: Employed and currently working (or on annual leave/holiday), Employed but on paid leave (including furlough), Employed and on unpaid leave, Apprenticeship, Self-employed and currently working (or on holiday)).

Non-employment (1: In unpaid/voluntary work, Self-employed but not currently working, Unemployed, Permanently sick or disabled, Looking after home or family, In education at school/college/university, Retired, Doing something else).

#### ELSA

Participants were asked two questions: “Which of these would you say best describes your current situation?” and “Are you currently looking for a job?” Based on responses to these questions participants were classified into “Economic activity” and “Employment status” as follows:

##### Economic activity

Economically active (0: Employed, Paid/unpaid leave from employment (including furlough), Self-employed and currently working, Self-employed but not currently working, Unemployed and looking for work).

Economically inactive (1: Retired, Unemployed and not looking for work, Permanently sick or disabled, Looking after home or family).

##### Employment status

Employed (0: Employed, Paid/unpaid leave from employment (including furlough), Self-employed and currently working, Self-employed but not currently working).

Non-employment (1: Retired, Self-employed but not currently working, Unemployed, Permanently sick or disabled, Looking after home or family).

#### Understanding Society

In each of the Covid waves participants were asked if their employment status had changed; if it had they were asked the follow-up question: “Even if you did not do any paid work last week, are you currently employed or self-employed?” In addition, in the January 2021 survey participants aged 65 years and under were asked: “Have you looked for any paid work or government training scheme in the last four weeks?” and “How many hours did you work, as an employee or self-employed, last week? Please include all jobs and self-employment activities. If you didn’t work any hours in your job(s) please enter zero”. These variables were used to derive Economic activity and Employment status variables as follows:

##### Economic activity

Economically active (0: Employed, Self-employed, Both employed and self-employed, and Not employed but looking for work).

Economically inactive (1: Not in employment and not looking for work).

##### Employment status

Employed (0: Employed, Self-employed and working more than 0 hours, Both employed and self-employed).

Non-employment (1: Not employed, Self-employed but working 0 hours).

### Appendix C: Coding self-reported measures of COVID-19

The self-reported measures of COVID-19 were derived using responses to interview questions for all waves up to and including the wave in which economic activity and employment status measures were recorded. Participants were defined as having had a test-confirmed case of COVID-19 if they reported in any wave that they had a positive test for COVID-19, and in the absence of a report of a positive test they were classified as a suspected case of COVID-19 if survey responses warranted it. The remaining participants were classified as not having had COVID-19. The wording for each of the cohorts is below.

#### CLS studies: BCS70, NCDS, NS

In each wave participants in the CLS studies were asked: “Do you think that you have or have had Coronavirus?”, with the responses: “Yes, confirmed by a positive test”, “Yes, based on strong personal suspicion or medical advice”; the remaining categories “Unsure” or “No” were considered to indicate not having had COVID-19.

#### ELSA

In both COVID-19 surveys ELSA participants were asked: “What was the result of your coronavirus (Covid-19) test? (If you had more than one test, please select ‘positive’ if any one of your tests have been positive).” The confirmed positive group consisted of those who reported a positive test, while those indicating a negative test were included in the “No COVID-19 category” In the first COVID-19 Survey participants were asked: “Since the coronavirus outbreak began in February, have you experienced any of the following symptoms of coronavirus (COVID-19)? After which participants were asked of 8 possible symptoms. Those who reported any of the symptoms (and had not reported a positive test) were included in the suspected COVID-19 category for that wave. Those who did not reported any symptoms were included in the “No COVID-19 category.” In the second COVID-19 Survey people were considered having suspected COVID-19 if the reason given for having had a COVID-19 test was having had Covid-19 symptoms, and they had not had a positive or negative result.

#### Understanding society

In each wave (with some minor variations in wording and response participants “were asked if they had been tested for coronavirus {since the last time you completed this survey on}”;they were then asked whether or not they had a positive test. An affirmative response in any wave was used to indicate a confirmed positive case of COVID-19. Those who indicated that they did not have a positive test were included among those who had not had COVID-19. In addition, participants who provided a positive response to the question: “Have you experienced symptoms that could be caused by coronavirus (COVID-19) {since the last time you completed this survey on..?”} were classified as having suspected COVID-19. Those who reported having had symptoms were considered to have suspicion of COVID-19 unless they indicated they had a negative test (see above). Those who did not have any symptoms of COVID-19 were classified in the “No COVID-19” category.

### Supplementary tables

#### Supplementary Table 1: Odds ratios for economic inactivity and not being in work for the UKLLC sample with COVID-19 status ascertained using linked NHS data

| Variable (reference category) |  | Economic activity | | | |  | Employment status | | | |
| --- | --- | --- | --- | --- | --- | --- | --- | --- | --- | --- |
|  |  | Odds Ratio | 95% CI | | p |  | Odds Ratio | 95% CI | | p |
| **COVID-19 positive test (No)** |  |  |  |  |  |  |  |  |  |  |
| Yes |  | 1.08 | 0.68 | 1.73 | 0.734 |  | 1.09 | 0.77 | 1.55 | 0.622 |
| **Age standardised** |  |  |  |  |  |  |  |  |  |  |
| Continuous term |  | 5.31 | 3.74 | 7.52 | <0.001 |  | 3.45 | 2.63 | 4.52 | <0.001 |
| Quadratic term |  | 2.05 | 1.48 | 2.83 | <0.001 |  | 1.79 | 1.41 | 2.28 | <0.001 |
| **Sex (Men)** |  |  |  |  |  |  |  |  |  |  |
| Women |  | 1.36 | 1.09 | 1.69 | 0.006 |  | 1.37 | 1.17 | 1.61 | <0.001 |
| **Household composition (Alone)** |  |  |  |  |  |  |  |  |  |  |
| Partner |  | 1.22 | 0.91 | 1.64 | 0.181 |  | 1.08 | 0.86 | 1.36 | 0.488 |
| Partner & children |  | 1.12 | 0.82 | 1.54 | 0.463 |  | 1.11 | 0.88 | 1.41 | 0.378 |
| No partner & children |  | 0.57 | 0.32 | 1.03 | 0.061 |  | 0.99 | 0.69 | 1.43 | 0.976 |
| Other person |  | 0.89 | 0.47 | 1.69 | 0.727 |  | 1.01 | 0.66 | 1.57 | 0.947 |
| **Ethnicity (White)** |  |  |  |  |  |  |  |  |  |  |
| Asian |  | 0.75 | 0.34 | 1.64 | 0.468 |  | 0.91 | 0.54 | 1.51 | 0.707 |
| Black |  | 0.22 | 0.03 | 1.58 | 0.131 |  | 1.45 | 0.74 | 2.84 | 0.273 |
| Mixed |  | 1.70 | 0.72 | 3.99 | 0.224 |  | 1.05 | 0.49 | 2.22 | 0.904 |
| Other |  | 2.61 | 0.89 | 7.64 | 0.080 |  | 2.29 | 0.98 | 5.39 | 0.057 |
| **Shielding (No)** |  |  |  |  |  |  |  |  |  |  |
| Yes |  | 0.98 | 0.62 | 1.52 | 0.913 |  | 1.04 | 0.74 | 1.44 | 0.840 |
| **Self-rated health (Excellent)** |  |  |  |  |  |  |  |  |  |  |
| Very Good |  | 0.94 | 0.68 | 1.30 | 0.712 |  | 1.04 | 0.81 | 1.33 | 0.781 |
| Good |  | 1.10 | 0.79 | 1.53 | 0.577 |  | 1.20 | 0.94 | 1.55 | 0.150 |
| Fair |  | 1.02 | 0.67 | 1.54 | 0.927 |  | 1.40 | 1.03 | 1.89 | 0.031 |
| Poor |  | 2.11 | 1.07 | 4.16 | 0.031 |  | 2.29 | 1.34 | 3.91 | <0.002 |
| **NS-SEC (Higher management and professional)** |  |  |  |  |  |  |  |  |  |  |
| Lower management and professional |  | 1.42 | 1.02 | 1.97 | 0.037 |  | 1.70 | 1.31 | 2.21 | <0.001 |
| Intermediate |  | 1.33 | 0.90 | 1.95 | 0.149 |  | 1.37 | 1.01 | 1.87 | 0.044 |
| Small employer |  | 0.84 | 0.52 | 1.36 | 0.472 |  | 4.11 | 3.03 | 5.58 | 0.001 |
| Lower supervisory & technical |  | 1.70 | 1.14 | 2.54 | 0.010 |  | 1.79 | 1.16 | 2.77 | 0.009 |
| Semi-routine |  | 1.21 | 0.73 | 1.99 | 0.456 |  | 1.94 | 1.40 | 2.69 | <0.001 |
| Routine |  | 1.21 | 0.73 | 1.99 | 0.456 |  | 2.83 | 1.99 | 4.02 | <0.001 |
| Unclassifiable |  | 1.29 | 0.74 | 2.25 | 0.375 |  | 2.72 | 1.81 | 4.08 | <0.001 |
| **Keyworker status (No)** |  |  |  |  |  |  |  |  |  |  |
| Yes |  | 0.39 | 0.31 | 0.49 | <0.001 |  | 0.28 | 0.23 | 0.34 | <0.001 |
| **Education (NVQ 4 or 5)** |  |  |  |  |  |  |  |  |  |  |
| NVQ 3 |  | 0.92 | 0.67 | 1.25 | 0.590 |  | 0.87 | 0.69 | 1.10 | 0.237 |
| NVQ 1 & 2 |  | 1.11 | 0.86 | 1.43 | 0.431 |  | 1.02 | 0.84 | 1.23 | 0.879 |
| None |  | 0.71 | 0.45 | 1.10 | 0.126 |  | 0.78 | 0.57 | 1.07 | 0.125 |
| Unclassifiable |  | 0.95 | 0.55 | 1.64 | 0.0867 |  | 1.12 | 0.77 | 1.62 | 0.557 |
| **Cohort (BCS70)** |  |  |  |  |  |  |  |  |  |  |
| ELSA |  | 1.01 | 0.59 | 1.75 | 0.959 |  | 0.64 | 0.43 | 0.94 | 0.023 |
| NCDS |  | 1.81 | 0.83 | 3.97 | 0.138 |  | 0.97 | 0.57 | 1.66 | 0.919 |
| Next Steps |  | 0.88 | 0.53 | 1.46 | 0.629 |  | 0.88 | 0.61 | 1.26 | 0.474 |
| USoc |  | 1.43 | 0.95 | 2.16 | 0.085 |  | 0.95 | 0.73 | 1.24 | 0.714 |
| Constant |  | 0.03 | 0.02 | 0.05 | <0.001 |  | 0.06 | 0.04 | 0.09 | <0.001 |

#### Supplementary Table 2: Odds ratios for economic inactivity and not being in work for the UKDS sample with self-reported COVID-19 status

| Variable (reference category) |  | Economic activity | | | |  | Employment status | | | |
| --- | --- | --- | --- | --- | --- | --- | --- | --- | --- | --- |
|  |  | Odds Ratio | 95% CI | | p |  | Odds Ratio | 95% CI | | p |
| **Self-reported COVID-19 status** |  |  |  |  |  |  |  |  |  |  |
| Suspected |  | 1.02 | 0.80 | 1.29 | 0.901 |  | 1.07 | 0.90 | 1.28 | 0.448 |
| Test confirmed |  | 1.01 | 0.70 | 1.44 | 0.972 |  | 1.03 | 0.79 | 1.35 | 0.810 |
| **Age (z-score)** |  |  |  |  |  |  |  |  |  |  |
| Continuous term |  | 2.39 | 2.08 | 2.74 | <0.001 |  | 1.93 | 1.73 | 2.15 | <0.001 |
| Quadratic term |  | 1.54 | 1.36 | 1.75 | <0.001 |  | 1.35 | 1.23 | 1.49 | <0.001 |
| **Sex (Men)** |  |  |  |  |  |  |  |  |  |  |
| Women |  | 1.29 | 1.09 | 1.54 | 0.003 |  | 1.34 | 1.18 | 1.52 | <0.001 |
| **Household composition (Alone)** |  |  |  |  |  |  |  |  |  |  |
| Partner |  | 1.42 | 1.12 | 1.80 | 0.004 |  | 1.23 | 1.03 | 1.47 | 0.025 |
| Partner & children |  | 1.19 | 0.92 | 1.54 | 0.175 |  | 1.10 | 0.92 | 1.33 | 0.300 |
| No partner & children |  | 0.82 | 0.54 | 1.26 | 0.376 |  | 1.00 | 0.75 | 1.34 | 0.993 |
| Other person |  | 0.92 | 0.55 | 1.51 | 0.731 |  | 1.20 | 0.86 | 1.68 | 0.275 |
| **Country (England)** |  |  |  |  |  |  |  |  |  |  |
| Scotland |  | 0.95 | 0.69 | 1.30 | 0.755 |  | 0.90 | 0.71 | 1.14 | 0.392 |
| Wales |  | 1.05 | 0.72 | 1.53 | 0.792 |  | 0.99 | 0.74 | 1.32 | 0.942 |
| Northern Ireland |  | 1.48 | 0.88 | 2.48 | 0.136 |  | 1.05 | 0.67 | 1.63 | 0.843 |
| Elsewhere |  | 0.79 | 0.39 | 1.60 | 0.510 |  | 1.10 | 0.71 | 1.70 | 0.669 |
| **Shielding (No)** |  |  |  |  |  |  |  |  |  |  |
| Yes |  | 1.08 | 0.76 | 1.54 | 0.657 |  | 1.09 | 0.84 | 1.43 | 0.512 |
| **Self-rated health (Excellent)** |  |  |  |  |  |  |  |  |  |  |
| Very Good |  | 0.99 | 0.76 | 1.29 | 0.963 |  | 0.94 | 0.78 | 1.14 | 0.519 |
| Good |  | 1.13 | 0.87 | 1.48 | 0.359 |  | 1.13 | 0.93 | 1.37 | 0.213 |
| Fair |  | 1.17 | 0.85 | 1.61 | 0.342 |  | 1.21 | 0.95 | 1.53 | 0.115 |
| Poor |  | 2.15 | 1.28 | 3.63 | 0.004 |  | 1.87 | 1.23 | 2.85 | 0.003 |
| **Keyworker (No)** |  |  |  |  |  |  |  |  |  |  |
| Yes |  | 0.37 | 0.31 | 0.45 | <0.001 |  | 0.25 | 0.22 | 0.29 | <0.001 |
| **NS-SEC (Higher management and professional)** |  |  |  |  |  |  |  |  |  |  |
| Lower management and professional |  | 1.53 | 1.17 | 2.01 | 0.002 |  | 1.70 | 1.38 | 2.09 | <0.001 |
| Intermediate |  | 1.59 | 1.17 | 2.16 | 0.003 |  | 1.52 | 1.20 | 1.93 | 0.001 |
| Small employer |  | 0.90 | 0.61 | 1.33 | 0.594 |  | 4.13 | 3.25 | 5.24 | <0.001 |
| Lower supervisory & technical |  | 1.07 | 0.66 | 1.73 | 0.794 |  | 1.51 | 1.06 | 2.13 | 0.021 |
| Semi-routine |  | 1.95 | 1.42 | 2.68 | 0.000 |  | 2.11 | 1.64 | 2.71 | <0.001 |
| Routine |  | 1.59 | 1.08 | 2.33 | 0.019 |  | 2.74 | 2.08 | 3.62 | <0.001 |
| Unclassifiable |  | 2.14 | 1.38 | 3.32 | 0.001 |  | 2.82 | 2.04 | 3.90 | <0.001 |
| **Education (NVQ 4 or 5)** |  |  |  |  |  |  |  |  |  |  |
| NVQ 3 |  | 1.14 | 0.89 | 1.45 | 0.293 |  | 0.94 | 0.78 | 1.14 | 0.542 |
| NVQ 1 & 2 |  | 1.01 | 0.82 | 1.25 | 0.905 |  | 0.98 | 0.84 | 1.14 | 0.784 |
| None |  | 0.85 | 0.62 | 1.17 | 0.330 |  | 0.85 | 0.67 | 1.08 | 0.191 |
| Unclassifiable |  | 0.81 | 0.58 | 1.13 | 0.218 |  | 1.01 | 0.80 | 1.28 | 0.910 |
| **Cohort (BCS70)** |  |  |  |  |  |  |  |  |  |  |
| ELSA |  | 0.93 | 0.60 | 1.43 | 0.735 |  | 0.65 | 0.48 | 0.88 | 0.005 |
| NCDS |  | 0.91 | 0.61 | 1.35 | 0.647 |  | 0.87 | 0.66 | 1.15 | 0.332 |
| Next Steps |  | 1.47 | 0.84 | 2.57 | 0.181 |  | 0.98 | 0.66 | 1.45 | 0.923 |
| USoc |  | 1.42 | 1.03 | 1.96 | 0.030 |  | 0.91 | 0.75 | 1.12 | 0.369 |
| Constant |  | 0.01 | 0.01 | 0.02 | <0.001 |  | 0.05 | 0.04 | 0.07 | <0.001 |

#### Supplementary Table 3: Cross-tabulation between participants with self-reported COVID-19 confirmed with a test and those testing positive for COVID-19 from NHS records

| Self-reported measures | English NHS records | |
| --- | --- | --- |
|  | No linked record of positive test (n) | Linked record of positive test (n) |
| No COVID-19 | 6,422 | 14 |
| Suspected COVID-19 | 1,154 | 10 |
| Confirmed COVID-19 with a positive test | 73 | 455 |

Table contains participants aged 25 to 65 years living in England with permission for data linkage in BCS70, NCDS, Next Steps, ELSA, and USoc.

### Supplementary figures

#### Supplementary Figure 1a: flow chart for derivation of UKLLC sample

Excluded incomplete covariate data

N = 1,203

Not in employment pre-pandemic

N = 10,399

Not eligible due to not being a participant in key surveys

N = 13,436

Employed pre-pandemic

N = 10,967

Resident outside England

N = 1,498

Aged 25 to 65 years during study

N = 21,366

Cohort members in Safehaven

N = 41,715

Participation in key surveys appropriate to each study

N = 28,279

Excluded incomplete covariate data

N = 92

Outside age range

N = 6,913

English only participants

N = 9,469

Final analytic sample

N = 8,174

Permission for linkage to English NHS data granted

N = 8,266

#### Supplementary Figure 1b: flow chart for derivation of UKDS sample

Not in employment pre-pandemic

N = 12,082

Not eligible due to not being a participant in key surveys

N = 82,129

Employed pre-pandemic

N = 14,048

Aged 25 to 65 years during study

N = 26,130

All participants in utilised data files

N = 116,084

Participation in key surveys appropriate to each study

N = 33,955

Excluded incomplete covariate data

N = 167

Outside age range

N = 7,825

Final analytic sample

N = 13,881
